# Determinants of psychological distress during the COVID-19 pandemic and the lockdown measures: a nationwide on-line survey in Greece and Cyprus

**DOI:** 10.1101/2020.10.25.20219006

**Authors:** Petros Galanis, Evangelia Andreadaki, Efrosyni Kleanthous, Anastasia Georgiadou, Efi Evangelou, Georgios Kallergis, Daphne Kaitelidou

## Abstract

A limited number of studies have investigated the distress during the COVID-19 pandemic and the lockdown measures in the general population. We studied psychological distress during the COVID-19 pandemic and the lockdown measures in the general population in Greece and Cyprus. Also, we investigated several demographic, clinical and job characteristics of the participants as possible determinants of distress. Data collection was performed during lockdown measures in Greece and Cyprus (from April 21^st^ to May 4^th^ 2020). All participants provided informed consent to participate in the study. We used the Impact of Event Scale-Revised (IES-R) to measure the psychological distress in response to the COVID-19 pandemic and lockdown measures. Seventeen point four percent of the participants had overall IES-R score from 24 to 32 indicating that posttraumatic stress disorder (PTSD) is a clinical concern, while 33.5% had overall IES-R score >32 indicating that PTSD is a probable diagnosis. Females, Cyprus residents, participants that live with elderly people or patients with a chronic disease in home, participants with a mental health disease or/and chronic disease, participants under pharmaceutical treatment, participants that lost their work due to the pandemic and participants that work in hospital experienced greater distress. Also, increased age and decreased educational level was associated with increased distress. Findings suggest that our sample experienced great distress and this distress was affected by several demographic, clinical and job characteristics. Appropriate interventions should be established in order to support psychologically high risk groups and decrease their distress.

## Introduction

In recent years, infectious disease outbreaks, epidemics and pandemics are becoming an increased peril for public health safety due to their exponentially growing in severity and frequency (Harvard Global Health Institute, 2018; Bloom and Cadarette, 2019). Novel coronavirus strain named SARS-CoV-2 that causes COVID-19 severe respiratory illness, emerged from the Wuhan city of Hubei province in China on late December 2019. World Health Organization (WHO) declared a world pandemic on 11^th^ March 2020, following the spread in 114 countries (Di Gennaro et al., 2020). As of 5 June 2020 over 200 countries have been affected, coronavirus cases have exceeded 6.5 millions, whilst attributable deaths have reached 387 thousands (WHO, 2020).

Studies have already shown that mitigation stringent measures along with the steep global uprise morbidity and mortality of the COVID-19 sparkle alarming psychological responses and distress in citizens (Moghanibashi-Mansourieh, 2020; Qiu et al., 2020; Wang et al., 2020a). The mental burden on the public caused by quarantine and the uncertainty abound with respect to a new virus, has been well documented and further recognized in the context of Ebola (O’Leary et al., 2018), SARS (Sim et al., 2010) and H_1_N_1_ outbreaks (Pfefferbaum et al., 2012). Also, posttraumatic stress disorder (PTSD) is a severe mental health condition and a common consequence of pandemics (Cénat et al., 2020; Dutheil et al., 2020). Although literature on those chronically precedence pandemics has been mixed over the predictors of distress (Brooks et al., 2020), the COVID-19-initiated distress seems to have been correlated with specific predictors. The eerily psychological strain from Covid-19, has been linked with both personal and sociodemographic characteristics. Female gender, young age, working outside your residency, high concern about juvenile family member contracting COVID-19, experiencing chronic medical problems or current physical symptoms, having a family member or someone in your broader social environment infected, as well as a history of stressful situations have been identified as predominant triggering factors of distress (Mazza et al., 2020; Wang et al., 2020b).

Response of Greek and Cyprus governments due to the COVID-19 pandemic was fast and rigorous. In particular, in Greece, the first case was reported on 26^th^ February 2020 and the Greek government reacted rapidly applying the early installation of social tracing apparatus for those who contacted COVID-19 cases, the compelling escalation of public health guidance and restrictive measures, up to the final lock down on 23^rd^ March (World Health System Response Monitor, 2020). In the same way, Cyprus, ahead of most European nations, initiated a fast response after the first two reported cases within its borders on 9^th^ March 2020. The country partially sealed its borders on March 14, followed by a total lockdown two days later and a complete shutdown to all air links by March 21 (World Health System Response Monitor, 2020). To the best of our knowledge, only one study in Europe (Mazza et al., 2020) has already investigated the psychological distress and associated factors during the COVID-19 pandemic. Thus, the aim of this study was to assess psychological distress during the COVID-19 pandemic and the lockdown measures in the general population in Greece and Cyprus. Also, we investigated a great number of demographic, clinical and job characteristics of the participants as possible determinants of psychological distress.

## Method

### Study design

A cross-sectional study was conducted in Greece and Cyprus. Data collection was performed during lockdown measures and in particular during April 21^st^ to May 4^th^ 2020.

We used the Greek version of the Impact of Event Scale-Revised (IES-R-Gr) (Mystakidou et al., 2007) to measure the distress in response to the COVID-19 pandemic and lockdown measures. Also, we collected data regarding the demographic, clinical and job characteristics of the participants. In particular, demographic, clinical and job characteristics included gender, age, family status, under-age children, educational level, living together with elderly people or patients with a chronic disease, living alone or with others, chronic disease, mental health disease, pharmaceutical treatment, vaccination for seasonal flu, working status, loss of work due to the pandemic, remote work from home due to the pandemic, work in hospital and daily contact with other people due to work.

We used google forms to create an anonymous online version of the IES-R and the data regarding the demographic, clinical and job characteristics of the participants. Distribution of the online questionnaire was performed through social media and e-mails and a snowball sampling strategy, focused on recruiting the general population living in Greece and Cyprus, was utilized in order to reach a large a number of participants. The online survey was first disseminated to social media and e-mails in Greece and Cyprus and all responders were encouraged to pass it on to others in order to increase the probability to achieve a representative sample of the general population. Thus, the online questionnaire was openly accessible to the general public nationwide in Greece and Cyprus. Also, the online questionnaire was accompanied by a cover letter with a full explanation of the study design and procedures and the participants’ right to complete the questionnaire and participate anonymously in the study. Thus, all participants provided informed consent to participate in the study. In addition, only adults over 18 years old were allowed to complete the questionnaire in order to avoid ethical issues with children. The National Bioethics Committee of Cyprus approved our study.

### Questionnaire

We used the IES-R that consisted of 22 items in three subscales: (a) the intrusion subscale with eight items related to intrusive thoughts, nightmares, intrusive feelings and imagery, and dissociative-like re-experiencing, (b) the avoidance subscale with eight items related to feelings, situations and ideas and (c) the hyperarousal subscale with six items related to anger, irritability, difficulty concentrating, hypervigilance and heightened startle. Each item is rated by the participants on a scale from 0 to 4 (0 = “not at all,” 1 = “a little bit,” 2 = “moderately,” 3 = “quite a bit,” and 4 = “extremely”) for the past seven days. We adjusted the IES-R in case of the COVID-19 pandemic, asking from the participants to indicate how distressing each difficulty has been for them during the past seven days with respect to COVID-19 pandemic and the lockdown measures. Reliability of the IES-R was assessed with Cronbach’s alpha coefficient.

There are two different approaches for the scoring of the IES-R: (a) calculate the mean of each subscale and the score for each subscale ranges from 0 to 4, while the maximum overall score ranges from 0 to 12 (Motlagh, 2010), (b) calculate the raw sum of answers in all items and the maximum overall score ranges from 0 to 88 (Weiss, 2004; Mystakidou et al., 2007; Weiss, 2007). According to the creator of the IES-R, individuals with maximum overall IES-R score of above 23 are of concern (Weiss, 2004; Weiss, 2007). In particular, PTSD is a clinical concern in individuals with IES-R score from 24 to 32 (Asukai et al., 2002), while PTSD is a probable diagnosis for individuals with IES-R score of above 32 (Creamer et al., 2003). Increased scores on the IES-R and the subscales are representative of greater distress and are associated with increased concern for PTSD and health and well-being consequences. We treated the IES-R score in all these ways in order to get more accurate results. Since there is a controversial regarding the validity of the cut-off points for the IES-R, we chose to use the overall IES-R score from 0 to 12 and the subscales scores as the dependent variables. Also, due to the extremely high correlation (r=0.99, p<0.001) between overall IES-R score on the scale from 0 to 12 and the scale from 0 to 88, we chose to use the overall IES-R score on the scale from 0 to 12 as the dependent variable in order to be in accordance with the subscales.

### Statistical analysis

Continuous variables are presented as mean, standard deviation, median, minimum value, and maximum value, while categorical variables are presented as numbers and percentages. We used the Kolmogorov-Smirnov test and normal Q-Q plots to found out that IES-R scores followed normal distribution.

Chi-square test, chi-square trend test and independent samples t-test were used to estimate differences between demographic and job characteristics of the participants according to the country of residence.

Demographic, clinical and job characteristics of the participants were the independent variables, while the IES-R scores were the dependent variables. Bivariate analyses between independent variables and the IES-R scores included independent samples t-test (comparison between the IES-R scores and a dichotomous variable), Pearson’s correlation coefficient (comparison between the IES-R scores and a continuous variable) and Spearman’s correlation coefficient (comparison between the IES-R scores and an ordinal variable). Variables that were significantly different (p<0.20) in bivariate analyses were entered into the backward stepwise multivariate linear regression analyses with the IES-R scores as the dependent variables. Criteria for entry and removal of variables were based on the likelihood ratio test, with entering and remove limits set at p<0.05 and p>0.10. Multivariate linear regression analysis was applied for the control of each potentially confounding of each statistically significant predictive factor to the others. We estimated adjusted coefficients beta with 95% confidence intervals and p-values. P-values < 0.05 were considered as statistically significant. Statistical analysis was performed with the Statistical Package for Social Sciences software (IBM Corp. Released 2012. IBM SPSS Statistics for Windows, Version 21.0. Armonk, NY: IBM Corp.).

## Results

### Demographic, clinical and job characteristics

The study population consisted of 3929 residents in Greece (n=2501) and Cyprus (n=1428). The demographic, clinical and job characteristics of the participants according to the country of residence are shown in Table 1. The mean age of the participants was 37 years and 17.1% lived with elderly people or patients with a chronic disease, 17.4% had a chronic disease, 6.7% had a mental health disease, 24.9% were under pharmaceutical treatment and 6.7% were vaccinated for seasonal flu. Eight-point two percent of the participants lost their work due to the pandemic, 27.1% worked remotely from home due to the pandemic and 57.3% were in daily contact with other people due to their work.

**Table 1.** Demographic, clinical and job characteristics of the participants according to the country of residence.

| Characteristics | Cypriot residents |  | Greek residents |  | Total |  | P-value |
| --- | --- | --- | --- | --- | --- | --- | --- |
|  | N | % | N | % | N | % |  |
| Gender |  |  |  |  |  |  | 0.5<sup>a</sup> |
| Females | 1050 | 73.5 | 1864 | 74.5 | 2914 | 74.2 |  |
| Males | 378 | 26.5 | 637 | 25.5 | 1015 | 25.8 |  |
| Age<sup>b</sup> | 36.9 | 9.1 | 37.0 | 10.0 | 37.0 | 9.7 | 0.8<sup>c</sup> |
| Family status |  |  |  |  |  |  | <0.001<sup>d</sup> |
| Singles | 395 | 27.7 | 995 | 39.8 | 1390 | 35.4 |  |
| Married/in cohabitation | 925 | 64.8 | 1318 | 52.7 | 2243 | 57.1 |  |
| Widowers | 7 | 0.5 | 21 | 0.8 | 28 | 0.7 |  |
| Divorced | 101 | 7.1 | 167 | 6.7 | 268 | 6.8 |  |
| Under-age children |  |  |  |  |  |  | <0.001<sup>a</sup> |
| No | 712 | 49.9 | 1596 | 63.8 | 2308 | 58.7 |  |
| Yes | 716 | 50.1 | 905 | 36.2 | 1621 | 41.3 |  |
| Higher educational level |  |  |  |  |  |  | 0.23<sup>d</sup> |
| Elementary school | 3 | 0.2 | 3 | 0.1 | 6 | 0.2 |  |
| Intermediate school | 19 | 1.3 | 18 | 0.7 | 37 | 0.9 |  |
| High school | 153 | 10.7 | 263 | 10.5 | 416 | 10.6 |  |
| Two years degree after high school | 222 | 15.5 | 349 | 14.0 | 571 | 14.5 |  |
| University degree | 422 | 29.6 | 1038 | 41.5 | 1460 | 37.2 |  |
| MSc degree | 559 | 39.1 | 719 | 29.7 | 1278 | 32.5 |  |
| PhD degree | 50 | 3.5 | 111 | 4.4 | 161 | 4.1 |  |
| Elderly people or patients with a chronic disease in home |  |  |  |  |  |  | **<0.001<sup>a</sup>** |
| No | 1246 | 87.3 | 2012 | 80.4 | 3258 | 82.9 |  |
| Yes | 182 | 12.7 | 489 | 19.6 | 671 | 17.1 |  |
| Living |  |  |  |  |  |  | **<0.001<sup>a</sup>** |
| With others | 1309 | 91.7 | 2196 | 87.8 | 3505 | 89.2 |  |
| Alone | 119 | 8.3 | 305 | 12.2 | 424 | 10.8 |  |
| Number of others in house<sup>b</sup> | 2.8 | 1.6 | 2.4 | 1.5 | 2.6 | 1.6 | **<0.001<sup>c</sup>** |
| Chronic disease |  |  |  |  |  |  | 0.14<sup>a</sup> |
| No | 1163 | 81.4 | 2083 | 83.3 | 3246 | 82.6 |  |
| Yes | 265 | 18.6 | 418 | 16.7 | 683 | 17.4 |  |
| Mental health disease |  |  |  |  |  |  | 0.19<sup>a</sup> |
| No | 1342 | 94.0 | 2323 | 92.9 | 3665 | 93.3 |  |
| Yes | 86 | 6.0 | 178 | 7.1 | 264 | 6.7 |  |
| Under pharmaceutical treatment |  |  |  |  |  |  | 0.23 |
| No | 1088 | 76.2 | 1863 | 74.5 | 2951 | 75.1 |  |
| Yes | 340 | 23.8 | 638 | 25.5 | 978 | 24.9 |  |
| Seasonal flu vaccine |  |  |  |  |  |  | **<0.001<sup>a</sup>** |
| No | 1256 | 88.0 | 1754 | 70.1 | 3665 | 93.3 |  |
| Yes | 172 | 12.0 | 747 | 29.9 | 264 | 6.7 |  |
| Working status |  |  |  |  |  |  | **<0.001<sup>a</sup>** |
| Non employees | 197 | 13.8 | 490 | 19.6 | 687 | 17.5 |  |
| Employees | 1231 | 86.2 | 2011 | 80.4 | 3242 | 82.5 |  |
| Loss of work due to the pandemic |  |  |  |  |  |  | 0.73<sup>a</sup> |
| No | 1133 | 92.0 | 1844 | 91.7 | 2977 | 91.8 |  |
| Yes | 98 | 8.0 | 167 | 8.3 | 265 | 8.2 |  |
| Remote work from home due to the pandemic |  |  |  |  |  |  | **<0.001<sup>a</sup>** |
| No | 847 | 69.0 | 1508 | 75.2 | 2355 | 72.9 |  |
| Yes | 380 | 31.0 | 496 | 24.8 | 876 | 27.1 |  |
| Work in hospital |  |  |  |  |  |  | 0.86<sup>a</sup> |
| No | 838 | 68.1 | 1375 | 68.4 | 2213 | 68.3 |  |
| Yes | 393 | 31.9 | 636 | 31.6 | 1029 | 31.7 |  |
| Daily contact with other people due to work |  |  |  |  |  |  | **<0.001<sup>a</sup>** |
| No | 570 | 47.4 | 772 | 39.8 | 1342 | 42.7 |  |
| Yes | 632 | 52.6 | 1170 | 60.2 | 1802 | 57.3 |  |

<sup>a</sup> chi-square test
<sup>b</sup> mean (standard deviation)
<sup>c</sup> independent samples t-test
<sup>d</sup> chi-square trend test

### Impact of Event Scale-Revised

Cronbach’s alphas for the intrusion, avoidance, and hyperarousal subscales were 0.89, 0.8 and 0.87 respectively indicating very good reliability of the IES-R.

Descriptive statistics for the IES-R according to the country of residence are shown in Table 2. Mean overall IES-R score (scale from 0 to 12) was 3.52, while regarding the subscales the highest score was for the avoidance subscale (1.36) and the lowest score for the hyperarousal scale (1.03). Overall IES-R score and subscales scores were higher for Cypriot residents than Greek residents indicating that Cypriot residents experienced greater distress.

**Table 2.**
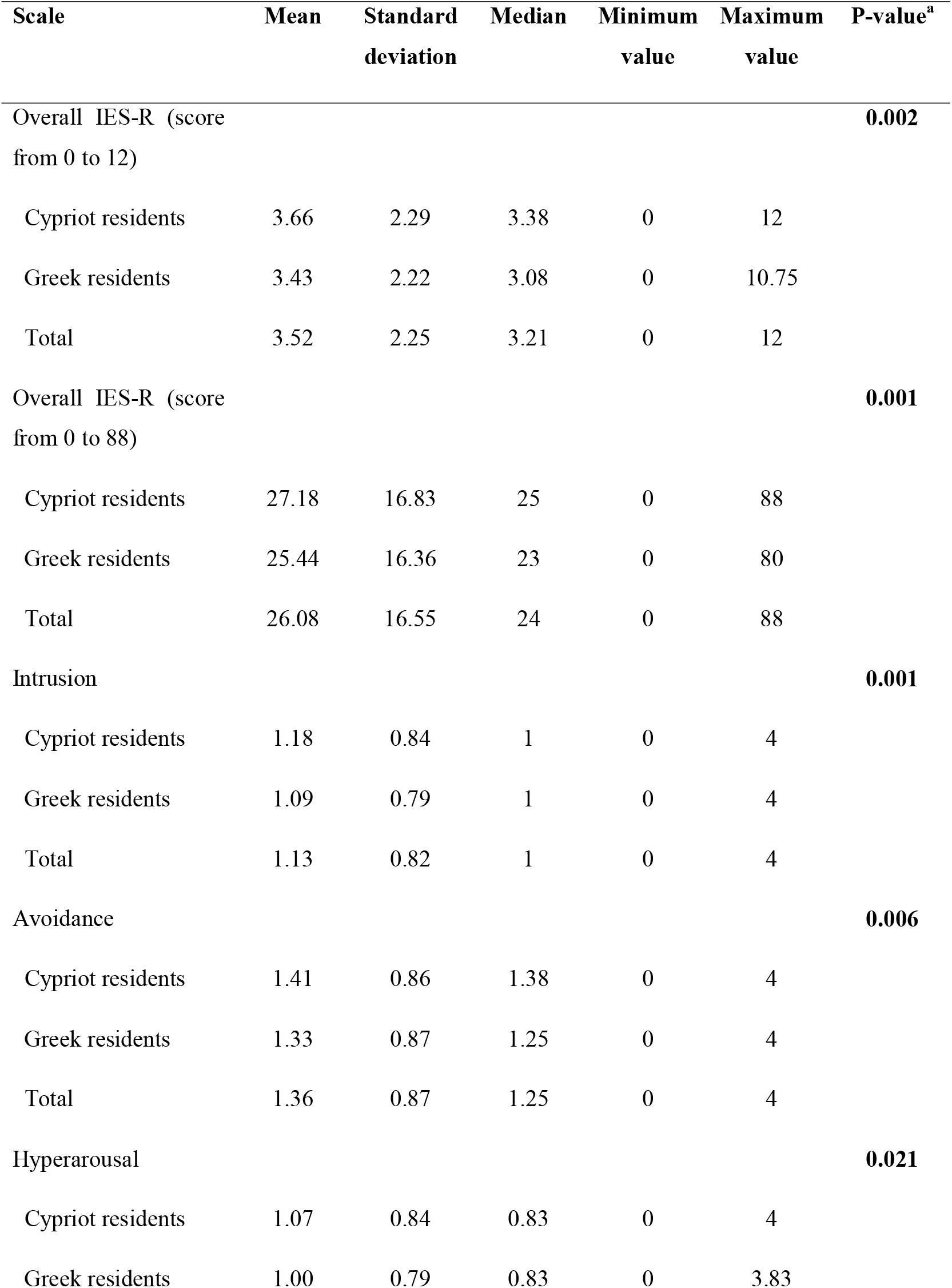
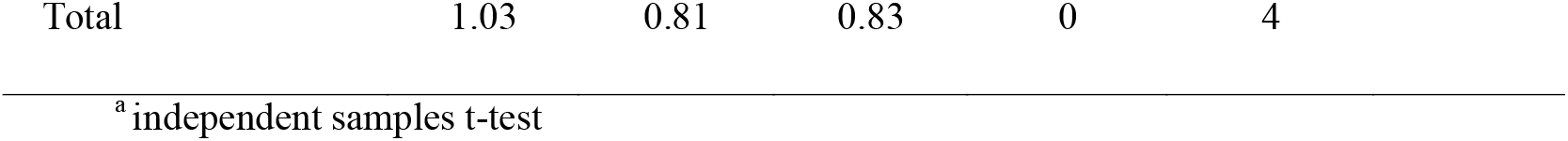
Descriptive statistics for the IES-R according to the country of residence.

| Scale | Mean | Standard deviation | Median | Minimum value | Maximum value | P-value<sup>a</sup> |
| --- | --- | --- | --- | --- | --- | --- |
| Overall IES-R (score from 0 to 12) |  |  |  |  |  | **0.002** |
| Cypriot residents | 3.66 | 2.29 | 3.38 | 0 | 12 |  |
| Greek residents | 3.43 | 2.22 | 3.08 | 0 | 10.75 |  |
| Total | 3.52 | 2.25 | 3.21 | 0 | 12 |  |
| Overall IES-R (score from 0 to 88) |  |  |  |  |  | **0.001** |
| Cypriot residents | 27.18 | 16.83 | 25 | 0 | 88 |  |
| Greek residents | 25.44 | 16.36 | 23 | 0 | 80 |  |
| Total | 26.08 | 16.55 | 24 | 0 | 88 |  |
| Intrusion |  |  |  |  |  | **0.001** |
| Cypriot residents | 1.18 | 0.84 | 1 | 0 | 4 |  |
| Greek residents | 1.09 | 0.79 | 1 | 0 | 4 |  |
| Total | 1.13 | 0.82 | 1 | 0 | 4 |  |
| Avoidance |  |  |  |  |  | **0.006** |
| Cypriot residents | 1.41 | 0.86 | 1.38 | 0 | 4 |  |
| Greek residents | 1.33 | 0.87 | 1.25 | 0 | 4 |  |
| Total | 1.36 | 0.87 | 1.25 | 0 | 4 |  |
| Hyperarousal |  |  |  |  |  | **0.021** |
| Cypriot residents | 1.07 | 0.84 | 0.83 | 0 | 4 |  |
| Greek residents | 1.00 | 0.79 | 0.83 | 0 | 3.83 |  |

|  |  |  |  |  |  |
| --- | --- | --- | --- | --- | --- |
| Total | 1.03 | 0.81 | 0.83 | 0 | 4 |

<sup>a</sup>independent samples t-test

Seventeen-point four percent of the participants (n=684) had overall IES-R score from 24 to 32 indicating that PTSD is a clinical concern, while 33.5% (n=1318) had overall IES-R score of above 32 indicating that PTSD is a probable diagnosis. Nineteen point three percent (n=275) of Cypriot residents had overall IES-R score from 24 to 32 and 34.9% (n=499) had overall IES-R score of above 32, while the respective percentages for the Greek residents were significantly lower; 16.4% (n=409) and 32.7% (n=819), (p=0.014).

### Determinants of distress

Bivariate analysis between independent variables (demographic and job characteristics of the participants) and IES-R scores are shown in Table 3, while multivariate linear regression analysis with IES-R scores as the dependent variables are shown in Table 4.

**Table 3.**
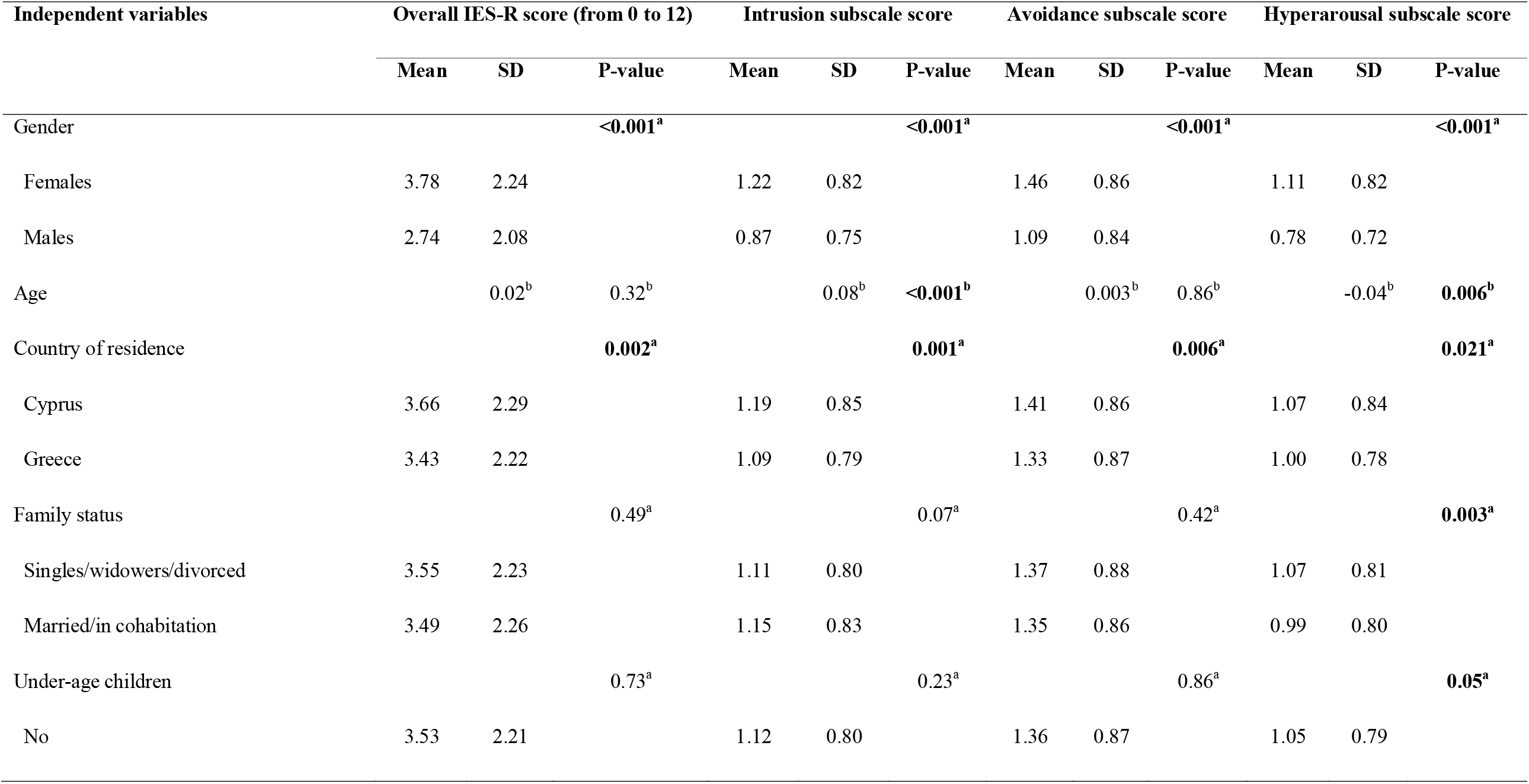
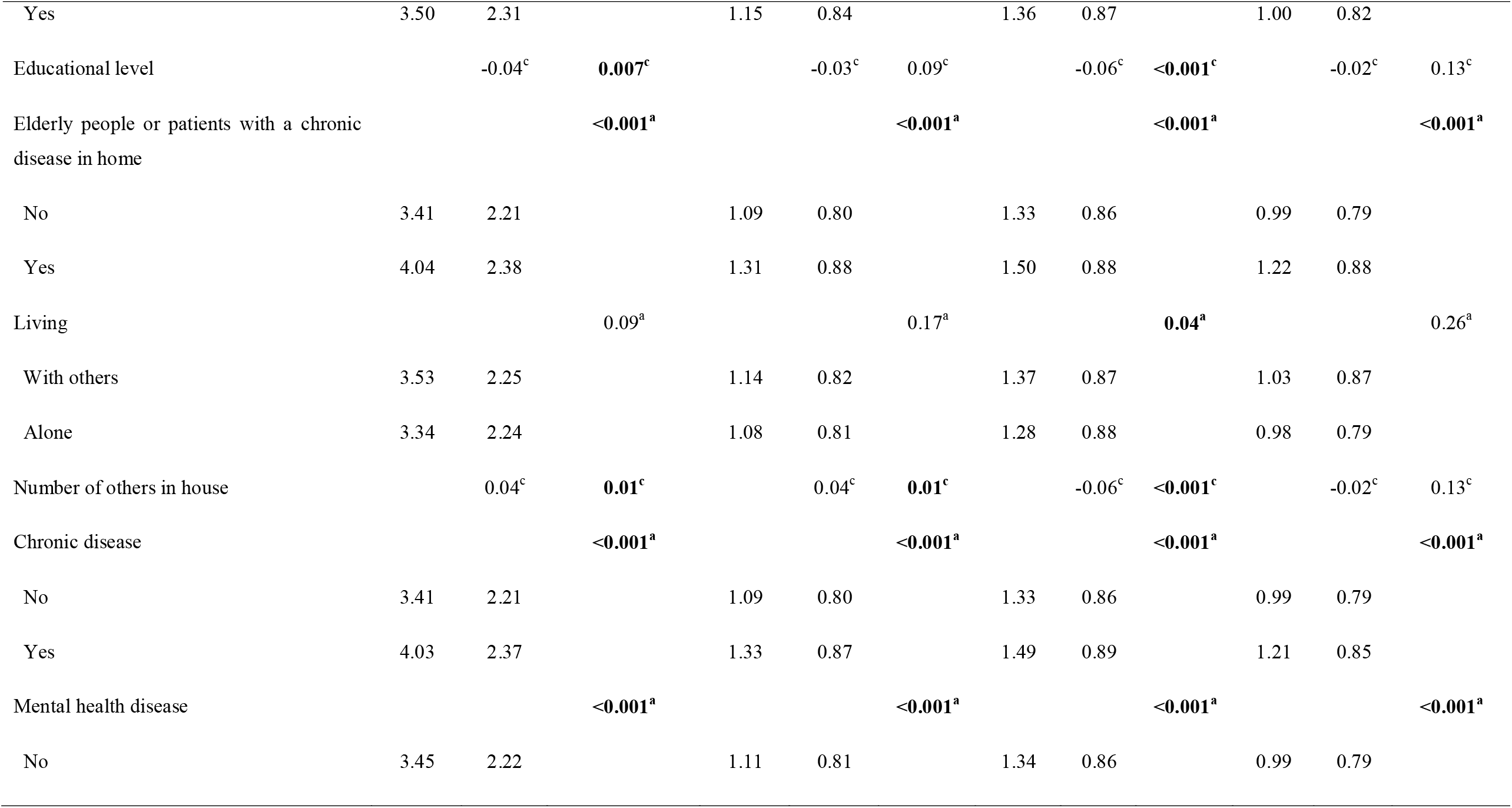
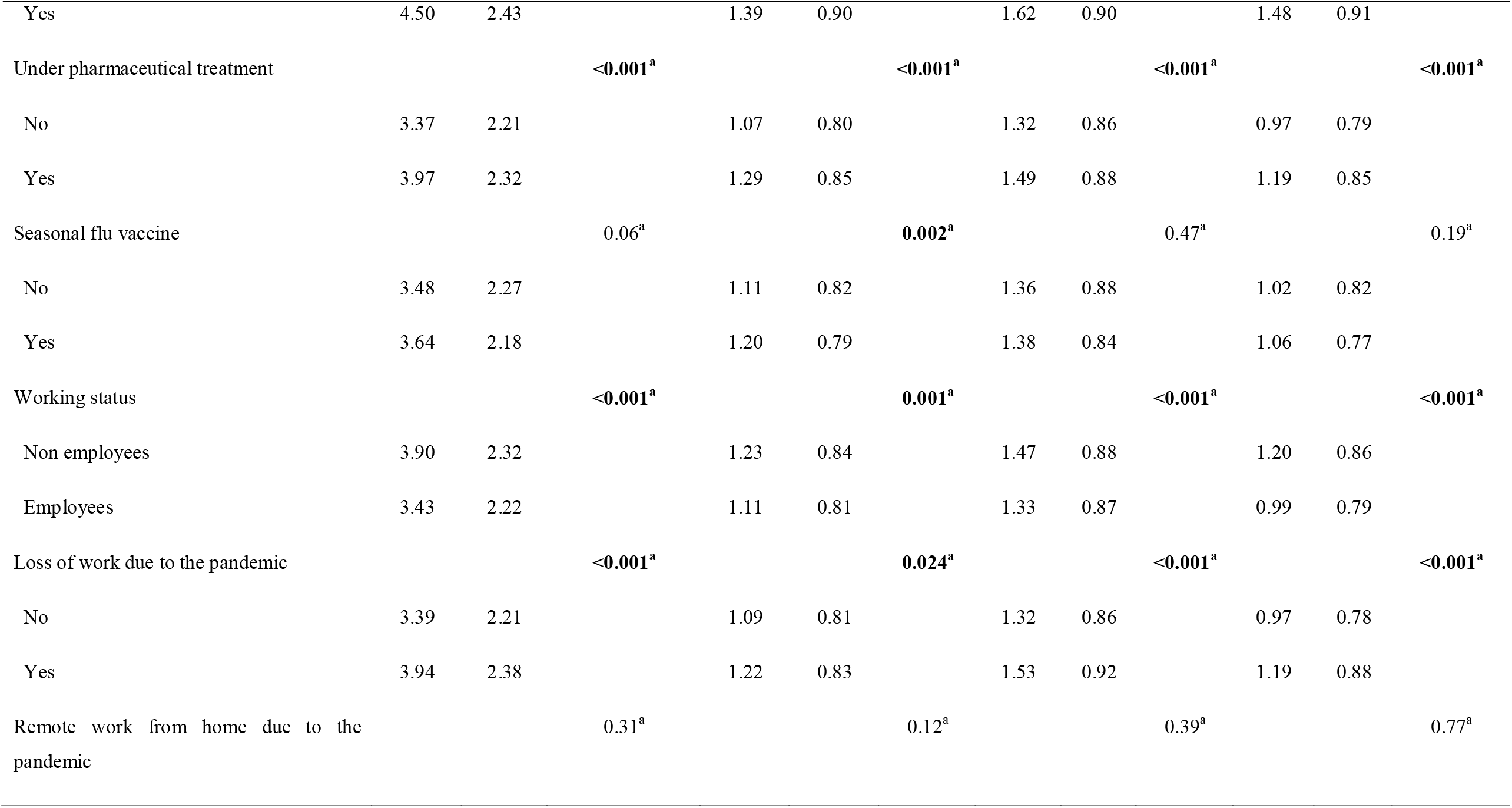
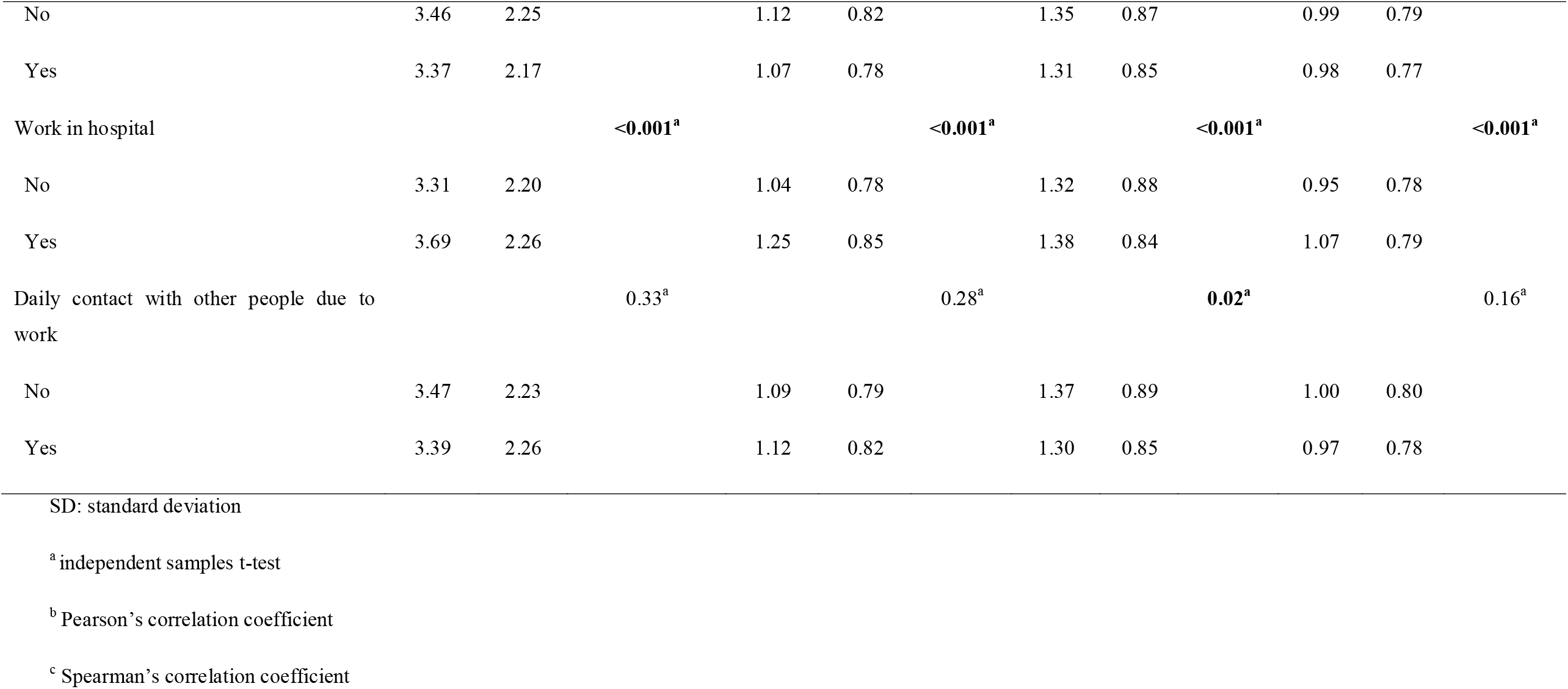
Bivariate analysis between independent variables and the IES-R scores.

**Table 4.**
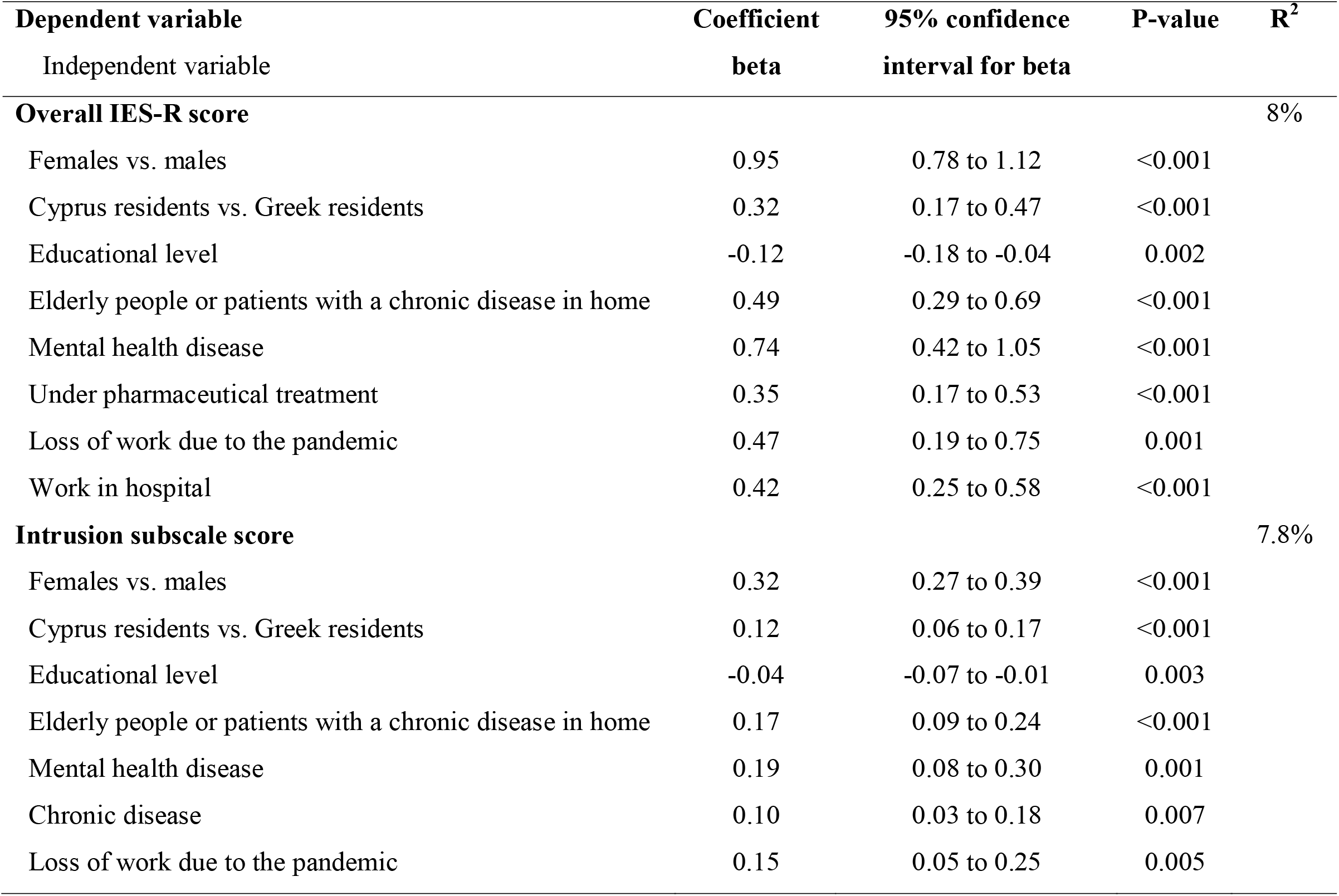
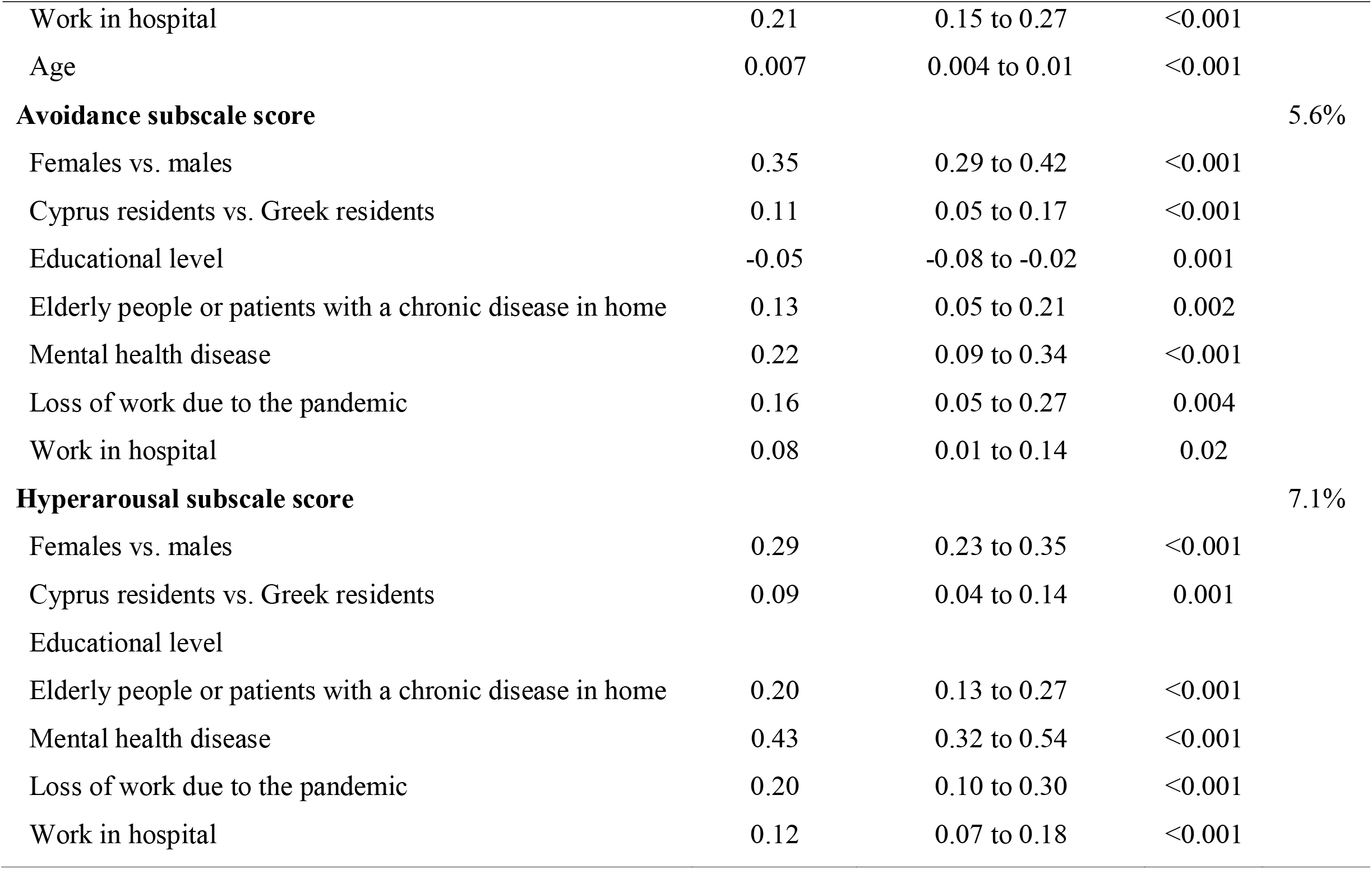
Multivariate linear regression analysis with IES-R scores as the dependent variables.

| **Dependent variable** | **Coefficient** | **95% confidence** | **P-value** | **R<sup>2</sup>** |
| --- | --- | --- | --- | --- |
| Independent variable | **beta** | **interval for beta** |  |  |
| **Overall IES-R score** |  |  |  | 8% |
| Females vs. males | 0.95 | 0.78 to 1.12 | <0.001 |  |
| Cyprus residents vs. Greek residents | 0.32 | 0.17 to 0.47 | <0.001 |  |
| Educational level | -0.12 | -0.18 to -0.04 | 0.002 |  |
| Elderly people or patients with a chronic disease in home | 0.49 | 0.29 to 0.69 | <0.001 |  |
| Mental health disease | 0.74 | 0.42 to 1.05 | <0.001 |  |
| Under pharmaceutical treatment | 0.35 | 0.17 to 0.53 | <0.001 |  |
| Loss of work due to the pandemic | 0.47 | 0.19 to 0.75 | 0.001 |  |
| Work in hospital | 0.42 | 0.25 to 0.58 | <0.001 |  |
| **Intrusion subscale score** |  |  |  | 7.8% |
| Females vs. males | 0.32 | 0.27 to 0.39 | <0.001 |  |
| Cyprus residents vs. Greek residents | 0.12 | 0.06 to 0.17 | <0.001 |  |
| Educational level | -0.04 | -0.07 to -0.01 | 0.003 |  |
| Elderly people or patients with a chronic disease in home | 0.17 | 0.09 to 0.24 | <0.001 |  |
| Mental health disease | 0.19 | 0.08 to 0.30 | 0.001 |  |
| Chronic disease | 0.10 | 0.03 to 0.18 | 0.007 |  |
| Loss of work due to the pandemic | 0.15 | 0.05 to 0.25 | 0.005 |  |
| Work in hospital | 0.21 | 0.15 to 0.27 | <0.001 |  |
| Age | 0.007 | 0.004 to 0.01 | <0.001 |  |
| **Avoidance subscale score** |  |  |  | 5.6% |
| Females vs. males | 0.35 | 0.29 to 0.42 | <0.001 |  |
| Cyprus residents vs. Greek residents | 0.11 | 0.05 to 0.17 | <0.001 |  |
| Educational level | -0.05 | -0.08 to -0.02 | 0.001 |  |
| Elderly people or patients with a chronic disease in home | 0.13 | 0.05 to 0.21 | 0.002 |  |
| Mental health disease | 0.22 | 0.09 to 0.34 | <0.001 |  |
| Loss of work due to the pandemic | 0.16 | 0.05 to 0.27 | 0.004 |  |
| Work in hospital | 0.08 | 0.01 to 0.14 | 0.02 |  |
| **Hyperarousal subscale score** |  |  |  | 7.1% |
| Females vs. males | 0.29 | 0.23 to 0.35 | <0.001 |  |
| Cyprus residents vs. Greek residents | 0.09 | 0.04 to 0.14 | 0.001 |  |
| Educational level |  |  |  |  |
| Elderly people or patients with a chronic disease in home | 0.20 | 0.13 to 0.27 | <0.001 |  |
| Mental health disease | 0.43 | 0.32 to 0.54 | <0.001 |  |
| Loss of work due to the pandemic | 0.20 | 0.10 to 0.30 | <0.001 |  |
| Work in hospital | 0.12 | 0.07 to 0.18 | <0.001 |  |

Females, Cyprus residents, participants that live with elderly people or patients with a chronic disease in home, participants with a mental health disease, participants that lost their work due to the pandemic and participants that work in hospital experienced greater distress according to overall IES-R score and the three subscales scores (intrusion, avoidance and hyperarousal). Also, decreased educational level was associated with increased distress in all IES-R scores. Participants under pharmaceutical treatment had higher overall IES-R score and participants with chronic disease had higher intrusion subscale score, while increased age was associated with increased intrusion subscale score.

## Discussion

Our nationwide study was among one of the first studies worldwide to investigate the distress during the COVID-19 pandemic and the lockdown measures. Also, we investigated several demographic, clinical and job determinants of distress in order to identify subgroups at greater risk of suffering from distress. To the best of our knowledge, only one study in Europe (Mazza et al., 2020) has already performed in this scientific area.

We found that PTSD due to the COVID-19 pandemic and the lockdown measures is a clinical concern in 17.4% of the participants and a probable diagnosis in 33.5%. In a similar study, Wang et al. used the same cut-off points with us regarding the IES-R and they found that PTSD is a clinical concern in 21.7% of the participants and a probable diagnosis in 53.8% (Wang et al., 2020a). Also, Zhang and Ma (2020) using a different cut-off of the IES ≥ 26 to reflect moderate to severe impact found that 7.6% of participants had an IES score ≥ 26. Mean overall IES-R score in our study was 26.1, while in Wang et al. (2020a) was 33 and in Zhang and Ma (2020) was 13.6. Also, a nationwide survey in China using a self-reported questionnaire found that almost 35% of the participants experienced psychological distress (Qiu et al., 2020). Thus, the COVID-19 pandemic and the lockdown measures have a moderate to severe impact in people’s life confirming that pandemics causes excessive anxiety, distress and panic. Additionally, quarantine during epidemics and pandemics increases the prevalence of psychological symptomatology e.g. stress, anxiety, depression and post-traumatic symptoms (DiGiovanni et al., 2004; Hawryluck et al., 2004; Jeong et al., 2016; Holmes et al., 2020; Brooks et al., 2020).

In our study, mental health disease was associated with greater distress. The COVID-19 epidemic has caused fear, anxiety, and depression. People with mental health conditions could be more substantially influenced by the emotional responses brought on by the COVID-19 epidemic, resulting in relapses or worsening of an already existing mental health condition because of high susceptibility to stress compared with the general population. A recently study in Italy found that patients with serious mental illness experienced higher levels of COVID-19-related perceived stress, anxiety, and depressive symptoms compared to control participants (Iasevoli et al., 2020). Many people with mental health disorders attend regular outpatient visits for evaluations and prescriptions. However, nationwide regulations on travel and quarantine have resulted in these regular visits becoming more difficult and impractical to attend (Holmes et al., 2020; Yao et al., 2020). In Greece and Cyprus, the majority of mental health services temporarily suspended their operation during the pandemic and offered only telephone support services. This may have increased the stress of the mentally ill.

Additionally, participants with chronic disease and those under pharmaceutical treatment experienced greater distress, a finding that is confirmed by the literature (Wang et al., 2020b). Comorbidity in patients with COVID-19 yielded poorer outcomes and was associated with substantial severity and mortality. In particular, cardiovascular diseases, cerebrovascular diseases, hypertension, malignancy, diabetes and chronic obstructive pulmonary disease seem to be the most significant risk factors for mortality from COVID-19 (Alqahtani et al., 2020; Du et al., 2020; Kumar et al., 2020; Guan et al., 2020). Thus, patients with chronic disease feel more vulnerable, anxious and depressed since they believe that they have an increased risk of death from COVID-19. In the same manner we could explain our finding that increased age of the participants was associated with increased distress. It is well known from the early studies regarding COVID-19 (Lai et al., 2020; Liu et al., 2020; Rothan and Byrareddy, 2020; Wu and McGoogan, 2020) that increased age is associated with increased mortality from COVID-19 and this information is widespread to the public resulting on increased anxiety and distress among elderly. Also, pre-existing depression in the elderly and their decreased access to mental health services increase COVID-19 related fear, distress and anxiety (Yang et al., 2020).

Also, we found that participants that work in hospital experienced greater distress. Medical staff was anxious regarding their safety and the safety of their families and reported psychological effects from reports of mortality from COVID-19 infection (Cai et al., 2020; Peeri et al., 2020). A study in China (Lai et al., 2020) revealed a high prevalence (71.5%) of mental health symptoms measured by the IES-R scale among health care workers treating patients with COVID-19, while a study (Reynolds et al., 2008) with persons quarantined during the SARS outbreak in Canada found that health care staff experienced greater psychological distress, including symptoms of PTSD. Also, Li et al., (2020) found that traumatization related to COVID-19 was higher among the general public than for front-line nurses. Tan et al., (2020) found that medical health care personnel experienced lower distress due to the COVID-19 pandemic than nonmedical health care personnel in two major tertiary institutions in Singapore. In the opposite, Chan and Huak (2004) found higher distress according to the IES score among physicians and nurses during the SARS outbreak, and an almost 3 times higher prevalence of PTSD, than this reported by Tan et al. (2020). This could be attributed to increased mental preparedness and stringent infection control measures after Singapore’s SARS experience.

Cyprus residents recorded a sobering preponderance of mental fallout in comparison to Greece residents and this finding can be in part attributed to the stringent governmental measures. In the interim of our study deployment, Cyprus residents already tallied more than a full month of total lockdown and communal life had been disrupted for a longer period of time. In Greece, measures taken where more gradual and reactive where, at a closer examination, that was a suitable line of action, considering that the attack rate of COVID-19 infection was significantly lower at all times in Greece. Indicative, on 21^st^ of April attack rate was 0.09 for Greece and 0.58 for Cyprus whereas by the end of this study, in Cyprus the rate had almost doubled, scoring 0.93, while on the other hand in Greece remained similar (0.06) (European Centre for Disease Prevention and Control, 2020). Taking that under account, as well as the fact that 14-day cumulative incidence of reported cases had been 35.52 and 4.56 of Cyprus and Greece respectively (World Health System Response Monitor, 2020) at the time our study kicked off, there are tangible evidence that COVID-19 had hit Cyprus residents way more belligerently than Greek community thus constituting a more stressful psychosocial cadre.

In our study, participants that lost their work due to the pandemic showed increased level of distress. The mental compressive load is even more exacerbated by the layoffs due to the pandemic-induced cease. Although pandemic-specific employment reinforcement incentives and unemployment benefits have been timely applied in both countries (International Labour organization, 2020; World Health System Response Monitor, 2020) as a state financial safeguard scheme, it has been shown that loss of income during the spread of communicable diseases, albeit relief policies, is a salient trigger of anxiety (Jeong et al., 2016). Even more, income reduction due to the SARS outbreak (Mihashi et al., 2009), was found to be a consequential risk factor for psychological disorders months after the acute phase had lapsed. Our finding that redundant workforce is shouldering a considerate heavier stress levels during pandemic might also come as a corollary to the loss of peer contact, deteriorating even more their social isolation.

In our study, participants that live with elderly people or patients with a chronic disease in home experienced greater distress. The importance of shielding the vulnerable population, such as the elderly and chronic diseases has been communicated throughout the media and through governmental public pleas repeatedly on numerous occasions (World Health System Response Monitor, 2020). The daily care of an elderly with disabilities is a stressful assignment of its own (Shen et al., 2019), on top of an impending infection menace that is highly dependent to physical distancing. Living with a vulnerable individual negates to effective distancing, and heightens the possibility of horizontal transmission. Our sample probably recognizing themselves as «Trojan horses» for SARS-CoV-2 to susceptible members of their family, recorded higher stress.

We found that being female was associated with greater distress. This finding is in line with the literature that female gender was associated with increased anxiety, depression, psychological distress and posttraumatic stress during epidemics and pandemics (Hawryluck et al., 2004; Lau et al., 2005; Mazza et al., 2020; Wang et al., 2020b). In general, females tend to be more vulnerable to experiencing stress and developing post-traumatic symptoms (Brewin et al., 2000; Norris et al., 2002).

### Limitations

Our study has several limitations. Given the limited resources available, quarantine restrictive measures and time-sensitivity of the COVID-19 pandemic, we collected the data through social media and a snowball sampling strategy was adopted, focused on recruiting the general population living in Greece and Cyprus. The snowballing sampling strategy is a quick and easy method but it is not a random procedure and thus generalization of our results should be done with great caution. Another limitation is that we used a self-reported questionnaire to measure the distress in response to the COVID-19 pandemic and lockdown measures and this measurement may not always be aligned with assessment by mental health professionals. Similarly, respondents might have provided socially desirable responses in terms of the satisfaction with the health information received and precautionary measures. In addition, we measured several demographic, clinical and job determinants of distress but there are many other determinants that could be investigated e.g. psychological support, financial status etc.

## Conclusions

Our findings mirror the trend in recent studies on the psychological impact of the COVID-19 pandemic among the general population. Identification of vulnerable populations and individuals at greater risk of suffering from psychological distress is a crucial step for immediate and effective actions to reduce the adverse psychological impact of the COVID-19 pandemic. For example, immediate and continuous psychological support is necessary in those with history of psychiatric illnesses as they have a greater likelihood of experiencing psychiatric symptoms. Since the impact of the COVID-19 pandemic could be long term, the health care system of countries should be focus on the mental health of individuals and especially of vulnerable populations. Also, tailored psychological interventions targeting the post-traumatic nature of the distress should be established especially for individuals at greater risk of suffering from psychological distress e.g. elderly, patients with mental disease, patients with chronic disease, medical and nursing staff etc. Further research is needed to explore the longitudinal mental health impact of the current pandemic especially to other vulnerable groups e.g. children, migrants, pregnant women, individuals with low socio-economic status etc.

## Data Availability

Availability of data and material: We can provide data (spss file).

## Notes

### Competing Interest Statement

The authors have declared no competing interest.

### Funding Statement

None to declare

### Author Declarations

Ethical Approval Reference Number: 2020.01.81 (National Bioethics Committee of Cyprus)

